# It’s not just about pads! Adolescent reproductive health in Kenya: *A qualitative secondary analysis*

**DOI:** 10.1101/2022.10.25.22281513

**Authors:** Sylvia Ayieko, Angela Nguku, Nancy Kidula

**Affiliations:** Department of Health Promotion and Behavioral Sciences, The University of Texas Health Science Center at Houston School of Public Health, Houston, Texas, United States of America; White Ribbon Alliance -Kenya, Nairobi, Kenya; White Ribbon Alliance-Global, Washington DC, United States of America; Department of Reproductive Health and Research (RHR), World Health Organization, Genève, Switzerland

**Author notes:** Corresponding Author: or (SA).

## Abstract

Many adolescents face barriers to accessing reproductive health care even though quality reproductive health care is a fundamental human right. This qualitative analysis describes quality reproductive health requests among adolescent high school girls in Kenya. We conducted a secondary analysis of qualitative data from a sub-sample of adolescent girls in Kenya who participated in the *What Women Want* global campaign. The campaign utilized one open-ended survey question. We also analyzed interview data from key informants involved in the survey. We used pre-existing codes and current literature to design the coding framework and thematic analysis to describe emerging themes. Atlas. ti 8 was used to organize and analyze codes. Over 4,500 high school girls, ages 12 and 19 years, were included in the analysis, with 61.6% from all-girls boarding schools and 13.8% from mixed-day schools. Data from nine key informants complemented findings from the survey. Emerging themes included: 1) The need for improved menstrual health and hygiene: Sanitary towels and cleaner toilets; 2) Prevention of adolescent pregnancy: Access to contraception; 3) Respect and dignity: Participants want privacy and confidentiality; and 4) The need to address social determinants of health: Economic stability and a safe physical environment. This study indicated that adolescent high school girls have varied requests for reproductive health care and services. While menstrual health and hygiene are key issues, reproductive needs are beyond just sanitary products. The results suggest a need for targeted reproductive health interventions using a multi-sectoral approach.

## Introduction

Access to health care, including sexual and reproductive health care, is a fundamental human right.^1,2^ However, many adolescents face barriers to accessing sexual and reproductive health services, which put them at risk for teenage pregnancy, unsafe abortions, reproductive tract cancers, and sexually transmitted infections, including Human Immunodeficiency Virus, Acquired Immunodeficiency Syndrome (HIV/AIDS).^3,4^ In Sub-Saharan Africa, for example, due to limited and inequitable sexual and reproductive health care services, adolescent girls and young women (15 to 24 years) bear a disproportionate burden (25%) of HIV infections, with new HIV infections occurring at a higher rate among 15–19-year-old females compared to their male counterparts.^5^

Pregnancy-related complications are among the leading causes of death among adolescent girls in Sub-Saharan Africa^6^ due to poor health care or non-utilization of health care services. Adolescent girls underutilize reproductive and maternal health services due to stigma, lack of information, weakened health systems, and poor implementation of health policies.^7,8^ In addition, poverty, low decision-making power, lack of negotiation skills, gender-based violence, and alcohol and drug abuse contribute to adolescents’ adverse reproductive health outcomes.^8,9^ In low-to-middle income countries, only about 10% of adolescent girls sought care from health facilities and received contraception counseling.^1^

In 2015, all 193 United Nations member states adopted the sustainable development goals (SDGs). Within the health-related SDG 3, targets 3·7 and 3·8 seek to ensure universal access to sexual and reproductive health care services, including family planning, information, and education, while striving to achieve universal health coverage, including access to quality essential health care services by 2030 respectively.^10,11^ *Reproductive health care for adolescents in Kenya*

Kenya, a lower-middle-income country in Eastern Africa, is a signatory to the SDGs and is committed to ensuring Universal Health Coverage. While the country has expanded access to adolescent sexual and reproductive health and rights (SRHR), impactful implementation of the SRHR interventions recommended by the *Global Accelerated Action for the Health of Adolescents (AA-HA!): Guidance to Support Country Implementation*^12^ has been limited. In 2017, about 24% of adolescent women aged 15 - 19 years in Kenya had an unmet need for family planning, resulting in 63% of unintended pregnancies in this population.^13^ Adolescent girls in Kenya are also more susceptible to HIV due to non-consensual, unprotected sexual intercourse and sexual violence.^14^ About 18% of teenage girls in Kenya between the ages of 15-19 years are either pregnant or mothers.^14^ Research suggests that factors associated with adolescent pregnancy in Kenya include lack of knowledge, unmet need for contraception, female genital mutilation (FGM) practices, and poverty.^15^ Adolescent girls in Kenya who engage in transactional sex due to low socioeconomic conditions also face challenges navigating through conflicting societal constructs that condemn adolescent contraceptive use and stigmatize pregnant adolescents.^16^

To reduce HIV infections and address unintended pregnancy in Kenya, the Kenyan government distributes free male condoms, targeting youth who may not afford or have access to condoms.^17^ Moreover, in collaboration with policymakers and civil society organizations, the Ministry of Health in Kenya has encouraged implementing the National Adolescent Sexual and Reproductive Health policy at all levels of care.^18^ The growing body of evidence suggests that the delivery of quality adolescent reproductive health care is linked to more positive health-seeking behaviors, increased adherence to birth control methods, and improved overall health outcomes among young people.^19^ Other studies allude to inadequate coordination and policy implementation that fails to incorporate pregnancy, menstruation, and sanitation into reproductive health.^20^ In addition, interventions on menstrual health hygiene programs in schools have not been well adopted because adolescents and young adults are not recognized as sources of information ^21,22^. Yet, they are the key stakeholders in their health.

Access to quality reproductive healthcare for adolescent girls in Kenya is a pressing public health issue warranting further investigation. To address this gap, this study utilizes qualitative data from the *What Women Want* survey, and interviews, conducted by the White Ribbon Alliance to explore what adolescent girls enrolled in different schools across Kenya demand as quality reproductive health care. It is expected that this secondary analysis of qualitative surveys and interviews will contribute to the current literature by discussing how adolescent girls conceptualize quality reproductive health and how to improve care.

## Materials and Methods

### Parent Study

The White Ribbon Alliance, a non-profit organization, conducted the global *What Women Want* campaign between April 2018 and March 2019 to understand what women and girls want for maternal and reproductive health care.^23,24^ The organization utilized a qualitative survey with one open-ended question: *My one request for quality reproductive and maternal health care services is ________?* Although the campaign initially administered the survey online, most countries, including Kenya, later distributed paper surveys and had different consenting procedures. In Kenya, data collection was via convenience sampling “citizen journalists,” community members, and volunteers associated with the White Ribbon Alliance in Kenya. About 120,000 women and girls completed the survey, with 16% being adolescents aged 15-19.^24^

### Ethics statement

De-identified data from the *What Women Want* results were obtained from the *What Women Want* dashboard.^24^ The current study does not involve interactions with human subjects but uses existing data. Given the nature of the study, the authors did not seek Institutional Review Board approval since the data was publicly available. The campaign outlined the consent processes during data collection for each country.^24^ In Kenya, verbal consent was obtained from girls and women who wished to remain anonymous and just provided their response and, in some cases, their age. In addition, approval and permission was obtained from schools before any data collection was done. Both verbal and written consent was obtained for adolescents aged 18 years or older who had their pictures taken.^24^

### Sample data source

We received authorization from the White Ribbon Alliance to conduct a secondary analysis of the qualitative survey from a sub-sample of adolescent girls in Kenya. The current study restricted analysis to data from teenage girls enrolled in sample high schools in eight counties in Kenya. Although some survey responses had missing information about the age or the type of school (due to the sensitive nature of the topic), the qualitative survey responses were sufficient for this current study. The study also included interview reports from nine key informants (teachers, youth advocates, and mobilizers) involved in the *What Women Want* campaign in Kenya.

### Type of schools

In Kenya, high schools are usually either day schools or boarding schools. Although few high (secondary) schools have some students enrolled as day scholars and others as boarders, this is not common. Moreover, boarding schools in Kenya are predominantly single-sex institutions. Thus, the survey responses from boarding schools reflected perspectives from mostly all-girls schools. Most day schools, by contrast, are co-ed/ mixed local schools, with many students making daily commutes from their homes.

### Data analysis

We conducted secondary analysis by re-examining survey responses from adolescent girls and analyzing transcripts from key informant interviews involved in the *What Women Want* campaign in Kenya. We conducted thematic content analysis based on emerging codes and existing quality-of-care frameworks^25^ to elucidate the phenomenon of quality reproductive care among adolescents. The initial coding system utilized categories and codes from the parent *What Women Want* study.^24^ SA conducted the initial coding by reviewing previously assigned codes and identifying mismatched responses. Then, three investigators (SA, AN, and NK) reviewed merging and reassigning codes. All the authors discussed the inconsistencies in the coding assignments and categories until the team achieved consensus. The iterative process resulted in new codes and categories for the coding framework. Thematic analysis was conducted using Atlas.ti-8 computer software.^26^

## Results

The secondary analysis included 4,570 survey responses from adolescent girls between 12 and 19 years in eight counties. Although 20% did not report their ages, their information was included for analysis. Most of the adolescent girls were enrolled in all-girls boarding schools (61·6%) compared to those who attended mixed-day schools (13·8%). **Table one** provides a summary of participant characteristics.

**Table 1:**
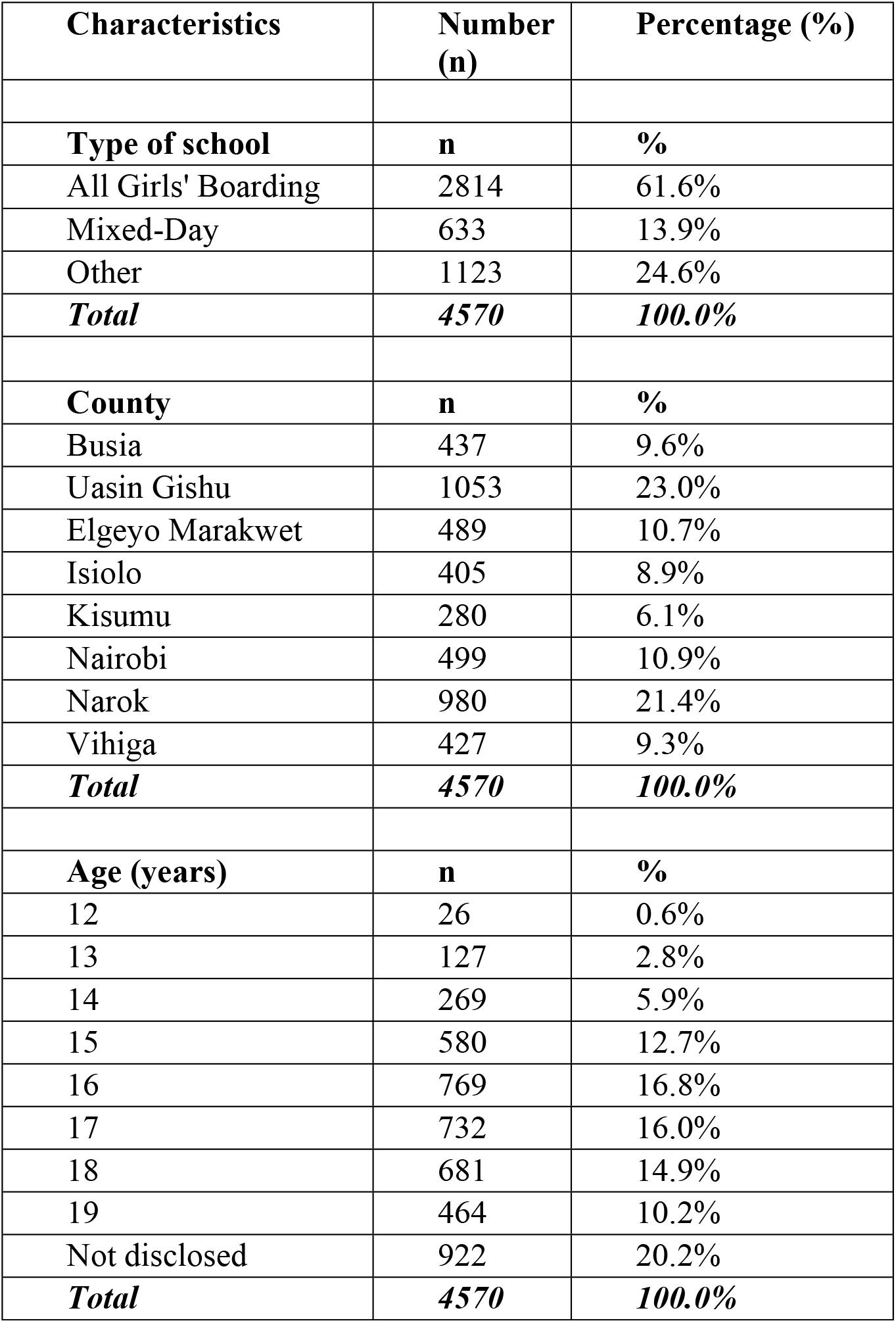
Demographic characteristics of study participants

| Characteristics | Number (n) | Percentage (%) |
| --- | --- | --- |
| **Type of school** | **n** | **%** |
| All Girls' Boarding | 2814 | 61.6% |
| Mixed-Day | 633 | 13.9% |
| Other | 1123 | 24.6% |
| **Total** | **4570** | **100.0%** |
| **County** | **n** | **%** |
| Busia | 437 | 9.6% |
| Uasin Gishu | 1053 | 23.0% |
| Elgeyo Marakwet | 489 | 10.7% |
| Isiolo | 405 | 8.9% |
| Kisumu | 280 | 6.1% |
| Nairobi | 499 | 10.9% |
| Narok | 980 | 21.4% |
| Vihiga | 427 | 9.3% |
| **Total** | **4570** | **100.0%** |
| **Age (years)** | **n** | **%** |
| 12 | 26 | 0.6% |
| 13 | 127 | 2.8% |
| 14 | 269 | 5.9% |
| 15 | 580 | 12.7% |
| 16 | 769 | 16.8% |
| 17 | 732 | 16.0% |
| 18 | 681 | 14.9% |
| 19 | 464 | 10.2% |
| Not disclosed | 922 | 20.2% |
| **Total** | **4570** | **100.0%** |

A few overarching themes emerged from the analysis of the findings from the qualitative survey and key informant interview data. The themes indicated the need for improved menstrual health and hygiene, adolescent pregnancy prevention, respect and dignity, and the need to address social determinants of health. **Table two** summarizes selected themes and examples of quotes from the qualitative survey.

**Table 2:** Sample categorization of themes, categories and codes used based on the survey responses.

| Themes | Categories | Codes | Examples |
| --- | --- | --- | --- |
| Improved Menstrual Health and Hygiene Management | Period poverty | Sanitary towels | *"Sanitary towels"* |
|  |  | Pain relief | *"Free sanitary towels for them to be comfortable during the periods and special treatment for those with cramps"* |
|  |  | Free sanitary products | *"The government to distribute pads to both primary and high schools, free Education on maternal health in different schools and institutions."* |
|  | Water and sanitation | Toilets/ Latrines | *"Improved toilets in schools that are hygienic"* |
|  |  | Water | *"Water in the toilets"* |
|  |  | Proper disposal | *"Teach women how to dispose of sanitary pads."* |
|  | Extra clothes and underwear | Extra skirts | *"Be given an extra skirt in case of periods leaking, which can cause embarrassment."*<br>*"I kindly request for underwear and bikers."* |
|  | Nutrition during menstruation |  | *"Give a list of \*nutritious foods, especially when it comes to monthly periods and cramps."* |
| Prevention of adolescent pregnancy | Risk Factors | Period poverty | *"Provide sanitary pads to the needy to reduce teenage pregnancy."* |
|  | Access to contraception | Condom use | *"Girls are becoming pregnant in our county at a young age; I would request the government to issue free condoms to the youth and \*educate girls about teenage pregnancy and its consequences."* |
|  |  | Abstinence | *"Teach the girls about abstinence from sex to avoid early pregnancy."* |
|  | Address cultural issues | Female genital mutilation | *"To prevent female genital mutilation and early pregnancy among girls to avoid school dropouts."* |
|  | Abortion | Prevention/ Care | *"Serious pregnancy tests to be conducted in institutions to prevent abortions."* |
| Respect and Dignity | Friendly | Non-abusive language | *"When young girls end up pregnant and go to a hospital to deliver, the nurses should be kind and stop using abusive languages since they lower self-esteem, most girls try to give birth on their own due to fear of bad treatment"* |
|  |  | Privacy/ confidentiality | *"A private and confidential room available concerning personal healthcare issues, provision of enough sanitary pads for quality education"* |
| Social determinants of health | Transportation |  | *"Provision of good road transport to and from school"* |
|  | Infrastructure | Roads | *"Government to improve local transport roads to enable easy access to the hospital."* |
|  | Poverty | Basic school needs | *"Boarding fees and uniform."* |
|  |  | Resources | *"Resources are targeted towards women with extreme poverty."* |

### Emerging themes

#### 1. Improved Menstrual Health and Hygiene

Menstrual health care encompassed the physical, emotional, and psychological elements and environmental factors such as the availability of water and toilets. The issues related to menstruation, such as sanitary products, safe disposal, the need for underwear, risk factors for period poverty, and pain medication, emerged across different ages, schools, and regions. One key informant stated:

> *“I engaged [with] young school-going girls above 15 years of age. Their main request was menstrual hygiene management and the provision of sanitary towels. What breaks my heart is how these goods do not reach some of these schools, and girls are missing school*.*” (Female Youth Advocate)*

Adolescent girls also expressed the need for water and sanitation during menstruation. Adolescent girls perceived clean toilets, water availability, and safe disposal of used menstrual products as quality reproductive healthcare. Responses ranged from needing more restrooms and accessible water sources to cleaner toilets and bathrooms. A few girls described unsanitary conditions as risk factors for reproductive health infections. One adolescent girl had this to say:

> *“To provide a private place where one can change or bath in case of messing because of her periods, have other places to dispose of *sanitary (sanitary towels) other than toilets (Adolescent girl, 17 yrs-Mixed day school)*.

In the analysis, the need for extra skirts, underwear, and other requests related to menstrual hygiene emerged as a significant ask among adolescent girls in mixed-day schools because they felt embarrassed when they stained their skirts during their period. A teacher in a mixed school in western Kenya further emphasized the theme of girls’ unique needs in her school.

> *“I buy the sanitary towels from my pocket because I care for the girls, but it is not enough. I also have extra skirts and keep a supply of pain relief medication. I have to have all these because we noticed that when the girls had their periods, they would skip school a few years back. Some were afraid of “spotting” their skirts and being laughed at by the boys, while others stated that they could not afford to buy sanitary pads*.*” (Female teacher - Mixed-day school)*

Many adolescent girls requested free or reduced costs of sanitary products and improved quality sanitary pads while directing their requests to government and non-profit organizations. There was also an emphasis of clean, safe spaces, indicating that adolescent girls in Kenya had different wants for menstrual health and hygiene.

#### 2. Prevention of Adolescent Pregnancy

Adolescents perceived prevention of adolescent/teenage pregnancy as quality reproductive care. Some categories associated with reduced adolescent pregnancies included pregnancy prevention methods, risk factors for adolescent pregnancy, abortion and post-abortion care, and proper care for pregnant teens and adolescent mothers. One mobilizer who interacted with teenagers reported that the girls were more concerned about contraception.

> *“For those who never had children, they were less concerned with what maternal health entails, but rather family planning affordability and accessibility” (Mobilizer, Nairobi)*

While some requested condoms and contraception pills for pregnancy prevention, others felt abstinence should be emphasized in schools. Some girls also criticized health care providers who prescribed family planning methods, stating that it promoted sexual immorality. Although the need for birth control access was prevalent, a few condemned oral contraception prescriptions, citing that it led to reproductive health issues.

A few girls mentioned the association between female genital mutilation, child marriages, and teen pregnancies. Other responses noted that the lack of sanitary pads puts girls at risk of teenage pregnancy and that providing menstrual supplies helps reduce teen pregnancies. Adolescents also had differing opinions about abortion. A few girls believed that conducting pregnancy tests in institutions could deter abortions, while others expressed the need for policies and legal processes around safe abortions. Some, however, felt that abortion was dangerous and should be condemned at all costs.

#### 3. Respect and dignity

Consistent with domains in quality-of-care frameworks, the request for respect and dignity was a key demand from adolescent girls. Responses indicated that adolescents want a safe space where their reproductive health needs are met without judgment. However, a few girls noted that health care workers treated pregnant teenagers and adolescent mothers disrespectfully as they sought care.

> *“When young girls end up pregnant and go to a hospital to deliver, the nurses should be kind and stop using abusive language since they lower self-esteem. Most girls try to give birth on their own due to fear of bad treatment” (Adolescent girl, 17 years-Boarding school)*

Adolescents perceived quality care as using appropriate language, with a few mentioning that they wanted doctors or nurses to be kind and polite and stop using abusive language. Some requested a change in attitude among providers and the elimination of harassment and criticisms that might lead to fear and low self-esteem among adolescent girls. A key informant, a teacher in an all-girls school, echoed the need for respectful care.

> *“Many girls quietly suffer because of reproductive health infections or sexually transmitted infections. Some who have contracted sexually transmitted infections rarely seek medical care for fear of being called out, especially when in line. Girls do not feel that the ‘youth-friendly corners’ meet their needs without prejudice*.*” (Female teacher-Boarding school)*

Many key informants noted that some teenage girls are reluctant to seek care because of negative past experiences with nurses.

#### 4. Addressing social determinants of health

Other themes emerged during the coding process that initially appeared unrelated to maternal or reproductive. In the parent study, some of these requests were classified as either “other asks” or “undeterminable asks.” However, based on literature and further examination, the authors considered economic situations, the physical environment, and social contexts as critical determinants of health that impact adolescent reproductive health outcomes. A youth advocate’s statement reinforces poverty’s impact on reproductive health.

“Don’t *make this just a pad issue!” Girls did not want pamphlets, materials, or instructions on menstrual health either. What they want, what they need are income-generating opportunities. “Girls told us it’s not just about whether they have sanitary napkins or not. It’s about the difficult choices between basic needs. “……. “If I’m comparing the price of sanitary towels or napkins to a packet of flour for dinner—I’m thinking I need to eat first*.*” (Female mobilizer-Nairobi)*

Access to food and nutrition was also linked to quality reproductive health care, with a few responses requesting balanced diets in schools and others demanding relief food in areas of famine. Some survey responses such as requests for “tuition and school uniform,” “need for books,” or “school fees” suggested that a few girls were from low socioeconomic households or families experiencing extreme poverty.

Participants not only expressed the need for economic empowerment but also felt that women should be actively involved in politics.

> *“Women should also participate actively in nation politics. This will help to fight for their rights freely and even come up with possible solutions to the problems that may be hindering them (17-year-old-Boarding school)*.

Adolescents also want improved physical environment for quality reproductive health care. For example, adolescents requested safe streets, proper lighting, and security in health facilities. Social inclusion emerged as a surprising sub-theme under social determinants of reproductive health. Adolescent girls conveyed the need to empower and educate women and girls to enhance their decision-making power.

## Discussion

Overall, the perceptions of quality reproductive care varied across adolescent high school girls in Kenya. Although the study results were similar to the overall *What Women Want* global survey findings, this analysis added value by highlighting unique themes among school-going adolescent females. Study findings acknowledge that while the survey question focused on maternal and reproductive healthcare services, the respondents also highlighted social determinants associated with the healthcare of women and girls. We identified issues adolescents felt were essential for quality reproductive health care.

A few emerging cross-cutting themes are consistent with adolescent reproductive health literature. For example, reducing teenage pregnancy rates is a health goal for many countries.^18, 27^ Engaging in transactional sex due to period poverty (lack of sanitary towels) is also a risk factor in other studies.^15,16^ Although the survey question was about requests for quality care rather than complaints, certain reproductive health care services, such as contraception, elicited negative feedback from some participants. Similar to other studies, there was evidence of stigma around adolescent use of contraception among a few respondents,^16^although many others requested increased access and additional information on birth control methods. The Adolescent Sexual and Reproductive Health policy in Kenya recognizes the need for a multi-sectoral approach in addressing teenage pregnancy,^18^ given the economic, sociocultural, and health system factors that impact adolescent girls.^15^ Historically, sociocultural systems prepared adolescents for menstruation, marriage, and pregnancy, where uncles and aunts provided sexuality education informally or during initiation ceremonies.^28^ However, given the societal shifts that emphasize nuclear families, and migration to cities for employment, adolescents have to rely on other sources for sexual and reproductive health education.

The need for improved menstrual health and hygiene was a common thread among female adolescents of all ages and schools. As such, we combined some categories (*water and sanitation* and *other determinants* “skirts”) into one theme, “Improved Menstrual Health and Hygiene,” based on the study findings. The request for additional skirts, including underwear, was more salient among girls in mixed-day schools probably because, similar to other studies, menstruating girls are likely to face ridicule from boys or community members when they spot during their period.^29^ Compared to girls in boarding schools who can go to their dorms during breaks and change, those in day schools are forced to continue classes the entire day with stained skirts or go home to change. It is also likely that some girls in day schools who come from poor households and cannot afford the cost of attending boarding schools also face challenges in obtaining sanitary products. Previous studies on menstruation report that absenteeism among adolescent girls was associated with menstruation due to a lack of sanitary towels, dysmenorrhea, embarrassment, or non-functional toilets.^29,30^ Although issues related to period poverty were significant requests for quality reproductive and maternal care, menstrual health management originated as an intervention related to education.^29^

The enactment of the Kenya Menstrual Hygiene Management Policy was lauded as a step toward improving educational outcomes for adolescent girls attending public schools in Kenya. However, there have been gaps in proper implementation due to challenges in the narrow conceptualization of menstrual health hygiene as just “pad distribution “.^31^ Studies in the United States and India also share similar findings on the need to frame Menstrual Health Hygiene as a comprehensive policy that impacts mental health, water and sanitation, education, and financial sectors.^31^As such implementation of menstrual health hygiene in schools not only addresses sustainable development goal (SDG) 3 of improving health and well-being but also SDG 4, in the provision of equitable and quality education especially for adolescent girls.^13, 14^

It is important to recognize that even though investments are made to improve sanitation and increase reproductive health supplies and products, adolescent girls may not fully seek care or utilize services if they feel disrespected and undervalued. Health care providers and practitioners working with adolescents should seek to uphold the dignity of adolescents without discrimination. Although it may be challenging, health providers should incorporate respectful language, dignified care, enhance privacy and confidentiality as a strategy for engaging adolescents seeking reproductive health care.

Overall, quality reproductive healthcare should not only be examined from the perspective of healthcare providers or professionals in the adolescent reproductive healthcare space. However, incorporating adolescent requests can be significant, as adolescents are experts on their health.^23^ The impact of economic and social determinants of health on reproductive health was also evident in the responses from adolescent girls in secondary schools in Kenya. As such, reproductive health programs should consider working with individuals in the ministries of Education, Finance, Food and Agriculture, Transport and Infrastructure, as well as those in the Sports, Culture & Heritage sectors.

## Strengths of the study

To our knowledge, this is the first study in Kenya on reproductive health that has asked women and girls what they want regarding quality reproductive health. By utilizing an open-ended question, participants felt free to express what they wanted and what they hoped could be implemented to improve their health. The large sample size elicited diverse responses on quality reproductive health care from the perspective of adolescent high school girls in Kenya. Along with these unique responses, our data highlighted the different demands by type of school and region, suggesting the need to target interventions unique to specific populations. The perspectives of key informants further reinforced the survey results. Reproductive health advocates are currently disseminating the findings from the study to policymakers and program implementers as evidence of demands from women and girls. Adolescent reproductive health care programs are likely to be more successful if they target the self-articulated needs of adolescents.

## Limitations

This study has limitations. First, the qualitative survey asked for only one request for quality healthcare. Adolescents probably had more requests but had to pick only one. Second, while the study offered girls a chance to speak up, it did not explore the reason behind the “asks” and how to navigate the issues. Additionally, this sub-sample was not nationally representative of all adolescent girls in Kenya. Some girls living with disabilities, in humanitarian settings, or in remote/ arid areas in Kenya may have different perceptions of quality reproductive health care. Future studies should explore conducting in-depth studies such as focus groups on quality reproductive healthcare among adolescent girls.

## Data Availability

Data from the main study can be obtained form the What Women Want dashboard: https://whiteribbonalliance.org/resources/www-dashboard/ Data will be made available by contacting the corresponding author.

https://whatwomenwant.whiteribbonalliance.org/

https://whiteribbonalliance.org/resources/www-dashboard/

## Acknowledgments

We would like to thank all the adolescent girls who completed the survey and their respective schools. We also appreciate the mobilizers, data collectors, and data entry personnel for their hard work and dedication. We would also like to acknowledge the White Ribbon Alliance staff who were involved in analyzing the entire global *What Women Want* survey.

## References

1. Woog V, Alyssa B, Jesse P. Adolescent Women’s Need for and Use of Sexual and Reproductive Health Services in Developing Countries. New York: Guttmacher Institute. 2015 [Cited 2022 August 9]. Available from: https://www.guttmacher.org/report/adolescent-womens-need-and-use-sexual-and-reproductive-health-services-developing-countries Accessed

2. World Health Organization: WHO recommendations on adolescent sexual and reproductive health and rights. [Cited 2022 August 9]. Available from: https://www.who.int/publications/i/item/9789241514606

3. Liang M, Simelane S, Fortuny Fillo G, et al. The State of Adolescent Sexual and Reproductive Health. J Adolesc Health. 2019;65(6S): S3–S15. doi:10.1016/j.jadohealth.2019.09.015

4. Ravindran TKS, Govender V. Sexual and reproductive health services in universal health coverage: a review of recent evidence from low-and middle-income countries. Sex Reprod Health Matters. 2020;28(2):1779632. doi:10.1080/26410397.2020.1779632

5. Murewanhema G. Adolescent girls, a forgotten population in resource-limited settings in the COVID-19 pandemic: implications for sexual and reproductive health outcomes. Pan Afr Med J. 2020;37(Suppl 1):41. doi:10.11604/pamj.supp.2020.37.41.26970

6. Grønvik T, Fossgard Sandøy I. Complications associated with adolescent childbearing in Sub-Saharan Africa: A systematic literature review and meta-analysis. PLoS One. 2018;13(9):e0204327. doi:10.1371/journal.pone.0204327

7. Hackett K, Lenters L, Vandermorris A, et al. How can engagement of adolescents in antenatal care be enhanced? Learning from the perspectives of young mothers in Ghana and Tanzania. BMC Pregnancy Childbirth. 2019;19(1):184. doi:10.1186/s12884-019-2326-3

8. Melesse DY, Mutua MK, Choudhury A, et al. Adolescent sexual and reproductive health in sub-Saharan Africa: who is left behind?. BMJ Glob Health. 2020;5(1):e002231. doi:10.1136/bmjgh-2019-002231

9. Birdthistle IJ, Floyd S, Machingura A, Mudziwapasi N, Gregson S, Glynn JR. From affected to infected? Orphanhood and HIV risk among female adolescents in urban Zimbabwe. AIDS. 2008;22(6):759–766. doi:10.1097/QAD.0b013e3282f4cac7

10. United Nations: Transforming our world: The 2030 agenda for sustainable development, United Nations A/RES/70/1, New York, 21 October 2015. [Cited 2022 August 9]. Available from: https://www.un.org/en/development/desa/population/migration/generalassembly/docs/globalcompact/A_RES_70_1_E.pdf

11. Waage J, Yap C, Bell S, et al. Governing the UN sustainable development goals: interactions, infrastructures, and institutions. Lancet Glob Health. 2015;3(5):e251–e252. doi:10.1016/S2214-109X(15)70112-9

12. World Health Organization. Global accelerated action for the health of adolescents (AA-HA!): guidance to support country implementation. Global Accelerated Action for the Health of Adolescents (AA-HA!) Guidance to Support Country Implementation. 2017. [Cited 2022 August 20]. Available from: https://www.who.int/publications/i/item/9789241512343

13. Darroch JE, Adding It Up: Investing in Contraception and Maternal and Newborn Health, 2017— Estimation Methodology, New York: Guttmacher Institute, 2018. [Cited 2022 August 20]. Available from: https://www.guttmacher.org/report/adding-it-up-investing-in-contraception-maternal-newborn-health-2017-methodology.

14. Kenya National Bureau of Statistics: Kenya Demographic Health Survey 2014. Nairobi, Kenya: Ministry of Health Kenya and ICF international. 2015. [Cited 2022 August 20]. Available from: https://dhsprogram.com/pubs/pdf/FR308/FR308.pdf.

15. Olenja J, Krugu JK, Kwaak v.d. A, Kawai D, Karanja S, Apanja M, et al., Factors influencing teenage pregnancy among Maasai girls in Kajiado West Sub-County, Kenya. An operational qualitative study report as part of the YES I DO programme implemented from 2016 to 2020. [Cited 2022 August 20]. Available from: https://www.kit.nl/wp-content/uploads/2021/01/YID-Kenya-Report-Teenage-pregnancy-final.pdf

16. Harrington EK, Casmir E, Kithao P, et al. “Spoiled” girls: Understanding social influences on adolescent contraceptive decision-making in Kenya. PLoS One. 2021;16(8):e0255954. doi:10.1371/journal.pone.0255954

17. Mburu A, Itsura P, Mabeya H, Kaaria A, Brown DR. Knowledge of Cervical Cancer and Acceptability of Prevention Strategies Among Human Papillomavirus-Vaccinated and Human Papillomavirus-Unvaccinated Adolescent Women in Eldoret, Kenya. Biores Open Access. 2019;8(1):139–145.. doi:10.1089/biores.2019.0007

18. Ministry of Health Kenya: Kenya Adolescent Sexual and Reproductive Health Policy. Nairobi, Kenya: Ministry of Health, Kenya, 2015. [Cited 2022 September 3]. Available from: https://www.popcouncil.org/uploads/pdfs/2015STEPUP_KenyaNationalAdolSRHPolicy.pdf

19. Mazur A, Brindis CD, Decker MJ. Assessing youth-friendly sexual and reproductive health services: a systematic review. BMC Health Serv Res. 2018;18(1):216. doi:10.1186/s12913-018-2982-4

20. Girod C, Ellis A, Andes KL, Freeman MC, Caruso BA. Physical, social, and political inequities constraining girls’ menstrual management at schools in informal settlements of Nairobi, Kenya. Journal of Urban Health. 2017 Dec;94(6):835–46. doi: 10.1007/s11524-017-0189-3.

21. Phillips-Howard PA, Caruso B, Torondel B, Zulaika G, Sahin M, Sommer M. Menstrual hygiene management among adolescent schoolgirls in low-and middle-income countries: research priorities. Global health action. 2016 Dec 1;9(1):33032. doi: 10.3402/gha.v9.33032.

22. Pilgrim NA, Jennings JM, Sanders R, et al. Understanding Quality of Care and Satisfaction With Sexual and Reproductive Healthcare Among Young Men. J Healthc Qual. 2018;40(6):354–366. doi:10.1097/JHQ.0000000000000149

23. Ayieko S, Nguku A. Engaging Youth in Citizen-Led Advocacy and Accountability for Adolescent Sexual and Reproductive Health. East Afr Health Res J. 2019;3(2):85–87. doi: 10.24248/eahrj.v3i2.603.

24. White Ribbon Alliance. What Women Want 2022. [Cited 2022 September 3]. Available from: https://whiteribbonalliance.org/resources/www-dashboard/

25. Jain AK, Hardee K. Revising the FP Quality of Care Framework in the Context of Rights-based Family Planning [published correction appears in Stud Fam Plann. 2018 Dec;49(4):397-401]. Stud Fam Plann. 2018;49(2):171–179. doi:10.1111/sifp.12052

26. ATLAS. ti Scientific Software Development GmbH. 2019. ATLAS. ti. [Cited 2022 September 3]. Available from: https://atlasti.com

27. Lugo-Gil J, Lee A, Vohra D, Harding J, Ochoa L, Goesling B. Updated findings from the HHS teen pregnancy prevention evidence review: August 2015 through October 2016. US Department of Health and Human Services. 2018. [Cited 2022 September 3]. Available from: https://tppevidencereview.youth.gov/pdfs/Summary_of_findings_2016-2017.pdf

28. De Haas B, Hutter I. Teachers’ conflicting cultural schemas of teaching comprehensive school-based sexuality education in Kampala, Uganda. Cult Health Sex. 2019;21(2):233–247. doi:10.1080/13691058.2018.1463455

29. Mohammed S, Larsen-Reindorf RE. Menstrual knowledge, sociocultural restrictions, and barriers to menstrual hygiene management in Ghana: Evidence from a multi-method survey among adolescent schoolgirls and schoolboys. PLoS One. 2020;15(10): e0241106. doi: 10.1371/journal.pone.0241106

30. Sommer M, Hirsch JS, Nathanson C, Parker RG. Comfortably, Safely, and Without Shame: Defining Menstrual Hygiene Management as a Public Health Issue. Am J Public Health. 2015;105(7):1302–1311. doi:10.2105/AJPH.2014.302525

31. Alhelou N, Kavattur PS, Rountree L, Winkler IT. ’We like things tangible:’ A critical analysis of menstrual hygiene and health policy-making in India, Kenya, Senegal and the United States [published online ahead of print, 2021 Dec 9]. Glob Public Health. 2021;1–14. doi:10.1080/17441692.2021.2011945

